# Age-dependent immune response to the Biontech/Pfizer BNT162b2 COVID-19 vaccination

**DOI:** 10.1101/2021.03.03.21251066

**Authors:** Lisa Müller, Marcel Andrée, Wiebke Moskorz, Ingo Drexler, Lara Walotka, Ramona Grothmann, Johannes Ptok, Jonas Hillebrandt, Anastasia Ritchie, Denise Rabl, Philipp Niklas Ostermann, Rebekka Robitzsch, Sandra Hauka, Andreas Walker, Christopher Menne, Ralf Grutza, Jörg Timm, Ortwin Adams, Heiner Schaal

## Abstract

**Background:** The SARS-CoV-2 pandemic has led to the development of various vaccines. Real-life data on immune responses elicited in the most vulnerable group of vaccinees over 80 years old is still underrepresented despite the prioritization of the elderly in vaccination campaigns.

**Methods:** We conducted a cohort study with two age groups, young vaccinees below the age of 60 and elderly vaccinees over the age of 80, to compare their antibody responses to the first and second dose of the BNT162b2 COVID-19 vaccination.

**Results:** While the majority of participants in both groups produced specific IgG antibody titers against SARS-CoV-2 spike protein, titers were significantly lower in elderly participants. Although the increment of antibody levels after the second immunization was higher in elderly participants, the absolute mean titer of this group remained lower than the <60 group. After the second vaccination, 31.3 % of the elderly had no detectable neutralizing antibodies in contrast to the younger group, in which only 2.2% had no detectable neutralizing antibodies.

**Conclusion:** Our data suggests that lower frequencies of neutralizing antibodies after BNT162b2 vaccination in the elderly population may require earlier revaccination to ensure strong immunity and protection against infection.

## Introduction

In December 2019, authorities in China’s Wuhan province reported a lung disease of unknown cause. Back in January 2020, the sequence of a novel coronavirus was published and identified as the causative agent of this disease [1]. In March of the same year, the World Health Organization (WHO) declared the spread of this virus a public health emergency of international concern. With limited drug treatment options available, research on prophylactic immunization, especially for high-risk groups, became a priority [2].

The zoonotic beta-coronavirus SARS-CoV-2 is closely related to severe acute respiratory syndrome coronavirus (SARS-CoV) and Middle East respiratory syndrome coronavirus (MERS-CoV), which caused outbreaks in 2002/2003 and 2012 respectively [3]. SARS-CoV-2 and its associated disease COVID-19, however, show distinct characteristics [4] including a highly variable severity of clinical symptoms, from asymptomatic infection to severe COVID-19 with lung manifestation and acute respiratory distress syndrome. Viral replication usually begins and continues in the upper respiratory tract for up to 14 days after the onset of symptoms, which contributes to the rapid spread of the virus. While the clinical course of COVID-19 is usually quite mild and often presents with flu-like symptoms, up to 14% of patients show a severe course of infection [5]. The elderly population is primarily at risk for severe disease, as adults over 65 years of age account for approximately 80% of hospitalizations [6, 7] and higher death rates have been reproducibly reported in this population [8, 9]. Additionally, prolonged disease from hospitalization, delayed viral clearance, and/or a higher fatality rate is also reported to be age-related [9]. Comorbidities such as cardiovascular disease, diabetes, and obesity are discussed as the primary cause of a more severe COVID-19 course, however, these comorbidities alone do not explain why age is such a strong risk factor.

Aging is accompanied by changes in the immune system, particularly affecting adaptive immunity’s three fundamental pillars, i.e. B cells, CD4+ T cells, and CD8+ T cells [10]. Although hallmarks of immunosenescence depend on multifaceted factors and vary greatly between individuals, they are generally considered to be related to i) the decreased ability to respond to new antigens associated with a reduced peripheral plasmablast response; (ii) decreased capacity of memory T cells and (iii) a low level of persistent chronic inflammation [11–14]. This leads to declining immune efficiency and fidelity, resulting in increased susceptibility to infectious diseases and decreased response to vaccinations. Additionally, it contributes to increased susceptibility to age-related pathological conditions including cardiovascular diseases or autoreactive diseases such as rheumatoid arthritis [12, 15].

In December 2020, the first vaccines for COVID-19 were approved worldwide and the first vaccinations were carried out [16–19]. While the German Standing Committee on Vaccination (STIKO) recommends immunization against SARS-CoV-2, access to the vaccine in Germany and many other countries worldwide at the beginning of 2021 is offered in a prioritization procedure due to limited availability. First, groups of people who are at particularly high risk for severe courses of COVID-19 disease or who are professionally in close contact with such vulnerable people were vaccinated. These two prioritized groups include senior residents of nursing homes aged ≥ 80 years, and their caregivers typically aged ≤ 65 years. A recent, thorough study using mathematical modeling to investigate vaccine prioritization strategies supports the preferential vaccination of the elderly [20]. This study describes a scenario where cumulative incidence rates were minimized when vaccination of the population aged 20-49 years was prioritized, while mortality was decreased when the population aged 60 years or older was prioritized. This model took age-structure, age-related efficacy, and infection-fatality rates into account. They conclude that prioritizing the population aged > 60 years, thus directly protecting the vulnerable population, would decrease mortality rates, a strategy that is currently employed by various nations but without the support of recent and thorough data [20].

The current vaccination strategy for the Biontech/Pfizer Comirnaty (BNT162b2) is a two-step “prime and boost” procedure in which the first vaccination is followed by a second vaccination with the same dose at least 21 days later [18]. Initial experience shows high effectiveness of the vaccines in preventing clinical symptoms after the first dose [21].

Immediately after the start of the official vaccination campaign in Germany at the end of December 2020, we started a daily practice study in a retirement home. To accommodate two clearly distinct populations in this study, we compared the induction of immune responses between young and elderly vaccinees (< 60 years of age and > 80 years of age respectively) who received their first and second vaccines on the same day. For this purpose, the IgG titers against SARS-CoV-2 spike S1 as well as neutralization titers were determined after both the first and second vaccination. Self-reported side effects were scored according to the sum of symptoms post-vaccination.

## Methods

### Study population

Characteristics of the study population are summarized in Table 1. The ethics committee of the Medical Faculty at the Heinrich-Heine University Düsseldorf, Germany (study no. 2021-1287), approved the study. Informed consent was obtained from all volunteers (N = 179) before sampling. All blood samples were collected on January 15^th^, 2021 (first collection, 17—19 days after first immunization) and February 5^th^, 2021 (second collection, 17 days after second immunization) and stored at 4 °C.

**Table 1:**
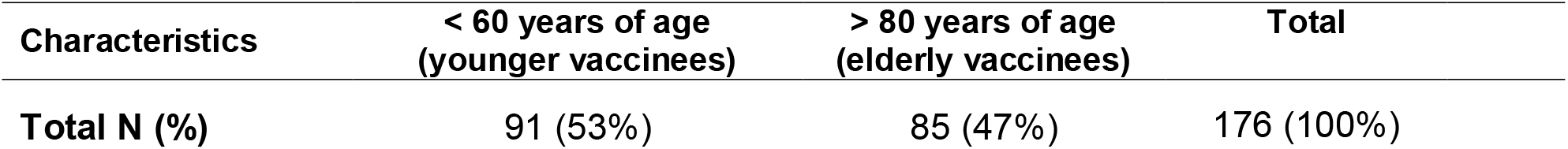
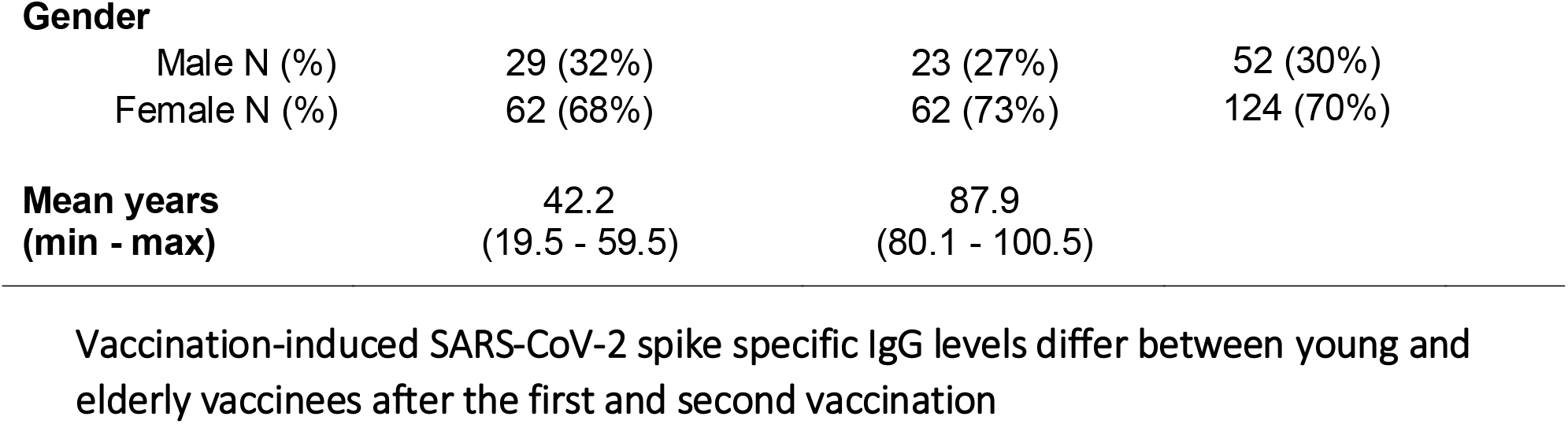
Characteristics of the study population

Medical questionnaires including the following categories were scored according to the sum of symptoms post-vaccination: i) elevated temperature and fever, ii) chills, iii) pain at the injection site, iv) head/limb pain, v) fatigue/tiredness, vi) nausea/dizziness, vii) other complaints (unscored).

### Commercially available Anti-SARS-CoV-2 tests systems

Samples were tested for Anti-SARS-CoV-2 antibodies using two commercially available test systems: Euroimmun Anti-SARS-CoV-2-QuantiVac-ELISA measuring IgG levels against SARS-CoV-2 spike S1 subunit and Abbott Architect SARS-CoV-2 IgG recognizing SARS-CoV-2 nucleocapsid (N) antibodies.

Euroimmun ELISA was performed on the Euroimmun Analyzer I-2P according to the manufacturer’s instructions. Results < 25.6 BAU/ml were considered as negative, ≥ 25.6 BAU/ml ≤ 35.2 BAU/ml as indeterminate, and > 35.2 BAU/ml as positive (BAU = Binding Antibody Units). The upper detection limit for undiluted samples was > 384 BAU/ml, the lower detection limit was < 3.2 BAU/ml. For samples over the detection limit, 1:10 or 1:100 dilutions were performed in IgG sample buffer according to the manufacturer’s instruction. The SARS-CoV-2 IgG chemiluminescent microparticle immunoassay (CMIA) from Abbott was performed on an ARCHITECT i2000 SR. The relation of chemiluminescent RLU and the calibrator is given as the calculated index (S/C). An index (S/C) <1.4 as was considered negative, ≥1.4 was considered positive.

### In-house SARS-CoV-2 neutralization test

A neutralization test with the infectious SARS-CoV-2 isolate (EPI_ISL_425126) was performed in a BSL-3 facility to determine the SARS-CoV-2 neutralization capacity of the serum samples after the first and second vaccination. A serial dilution endpoint neutralization test for SARS-CoV-2 was performed as previously described [22]. Serial dilutions of heat-inactivated (56°C, 30 minutes) serum samples were pre-incubated in cell-free plates with 100 TCID50 units of SARS-CoV-2 for 1 hour at 37° C. After pre-incubation, 100µl of cell suspension containing 7×10^4^/ml Vero cells (ATTC-CCL-81) were added. Plates were incubated at 37°C, 5% CO2 for 4 days before microscopic inspection for virus-induced cytopathic effect (CPE). The neutralization titer was determined as the highest serum dilution without CPE. Tests were performed in duplicate for each sample. Positive, negative, virus only, and cell growth controls were run during each assay.

### Statistical analysis

The data were analyzed using SPSS Statistics 25 (IBM^©^) and GraphPad Prism 9.0.00 (GraphPad Software, San Diego, CA, USA). Categorical data were studied using Fisher’s exact test or Pearson’s chi-square test, depending on the sample size. Quantitative data were analyzed by the non-parametric Mann-Whitney U test for two groups of paired and unpaired samples. Simple linear regression was performed using GraphPad Prism version 9.0.0 (the coefficient of determination R^2^ and p-values are given in the figures).

## Results

### Participant characteristics

In total, blood samples from 176 volunteers, young and elderly vaccinees (<60 / >80 years of age) were analyzed for vaccine-induced SARS-CoV-2 spike specific IgG titers and SARS-CoV-2 neutralizing antibodies after a prime and boost vaccination campaign using BNT162b2 (Comirnaty BioNTech/Pfizer) to screen for age-related differences in their immune response. Therefore, samples were collected at two time points, 17—19 days after the first vaccination and 17 days after the second vaccination. To be able to distinguish the immune response of the vaccinees from those who had already undergone a previous SARS-CoV-2 infection we also determined infection-induced SARS-CoV-2 nucleocapsid specific antibodies using the SARS-CoV-2 IgG chemiluminescent microparticle immunoassay (CMIA). Three vaccines were tested positive and therefore were excluded from the dataset. While group sizes were comparable (93 participants <60 years of age versus 83 participants >80 years of age), there was an overrepresentation of female participants compared with males (124 female to 52 male) (Table 1).

### Vaccination-induced SARS-CoV-2 spike specific IgG levels differ between young and elderly vaccinees after the first and second vaccination

The first sample collection was carried out 17—19 days after the volunteers received their first vaccination in late December 2020. At this timepoint, quantitative SARS-CoV-2 spike S1 specific IgG levels between the two groups differed significantly (p < 0.0001). For the younger group of vaccinees, IgG titers ranged between 0—3840.0 BAU/ml with a mean of 313.3 BAU/ml after the first vaccination. Only 4.4 % of the participants had titers below the cut-off, and 2.3% were indeterminate (Figure 1A). The mean titer for the group > 80 years of age was 41.2 BAU/ml with titers ranging from 0—484.7 BAU/ml. In this group, 65.9% showed titers below the cut-off (>35.6), and 9.4% were indeterminate.

**Figure 1.**
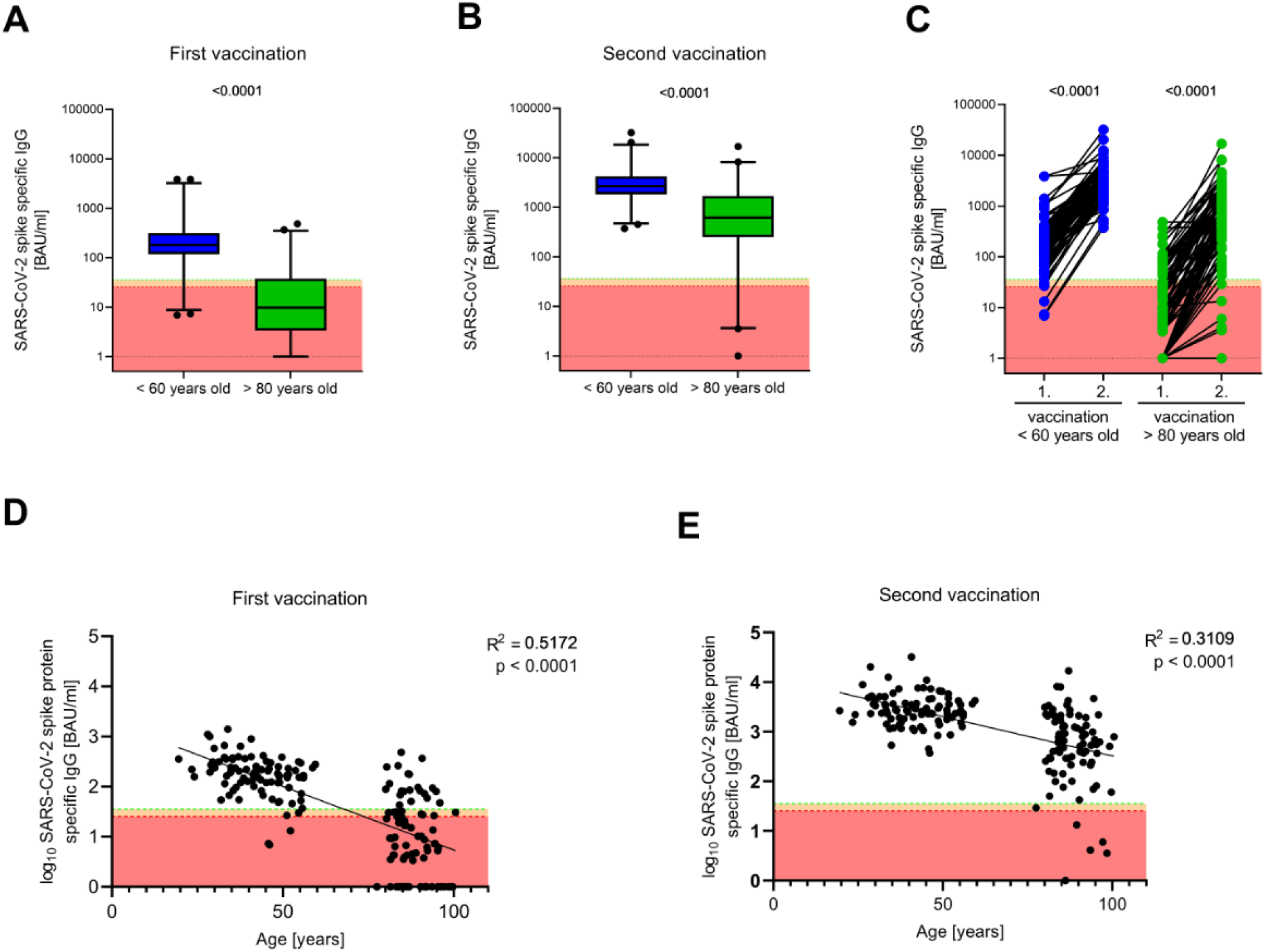
SARS-CoV-2 spike protein specific antibody titers were determined using Euroimmun Anti-SARS-CoV-2-QuantiVac-ELISA. Antibody titers below the detection limit were set to 1.0. **A** and **B** Antibody titers 17—19 day after first (A) and second (B) vaccination are shown. Boxes span the interquartile range; the line within each box denotes the median and whiskers indicate the 2.5 and 97.5 percentile values. **C** The pairwise comparison of IgG antibody titers within the two analysed age groups are shown. **D** and **E** Linear correlations between participant’s age and SARS-CoV-2 specific antibody titer after first vaccination (D) and second vaccination (E). Results < 25.6 BAU/ml as negative (red area), ≥ 25.6 BAU/ml ≤ 35.6 BAU/ml as indeterminate (orange), and > 35.6 BAU/ml were considered positive. For comparison of two groups either two-tailed parametric unpaired t-tests or paired t-test were performed. Correlation was analysed by simple linear regression. P-values < 0.05 were considered statistically significant. P-Values are depicted in the figures.

The second sample collection was carried out 17 days after the volunteers received their second vaccination, at a time point when full protection is suggested (>7 days according to [18]). Nevertheless, there was still a significant difference in IgG levels between the two groups. The mean titer of the younger group increased more than 10-fold (3702.0 BAU/ml) and ranged from 81.6—32000.0 with no participant testing below cut-off (Figure 1B). While the mean titer for elderly vaccinees increased to 1332.0 BAU/ml (0—16891.0 BAU/ml), 10.6% of the participants in this group still had titers below the cut-off.

The comparison of SARS-CoV-2 spike specific IgG titers showed an extremely significant (p<0.001) difference between the two age groups, after both the first and second vaccination, suggesting an attenuated antibody response in the group of elderly vaccinees > 80 years of age. While the gap in mean values narrowed after the second vaccination, which in particular underlines once again the necessity of a second vaccination, several elderly participants remained below the detection limit of the anti-SARS-CoV-2 assay. A general age-dependent negative correlation in SARS-CoV-2 spike specific IgG after both vaccinations is noticed throughout the entire cohort (Figure 1D/1E).

### Elderly vaccinees showed reduced SARS-CoV-2 neutralizing capacity compared to younger vaccinees

We next determined the neutralization capacity in our cohort after the first and second dose of vaccination. At 17—19 days after the first vaccination, the majority of participants, regardless of their age, failed to display neutralizing antibody titers. In the group of younger vaccines, 16.1 % displayed neutralizing antibodies with titers ranging between 1:10 to 1:2560. In the group of elderly vaccinees, only 1.2 % had developed neutralizing antibodies after the first vaccination (Figure 2A).

**Figure 2.**
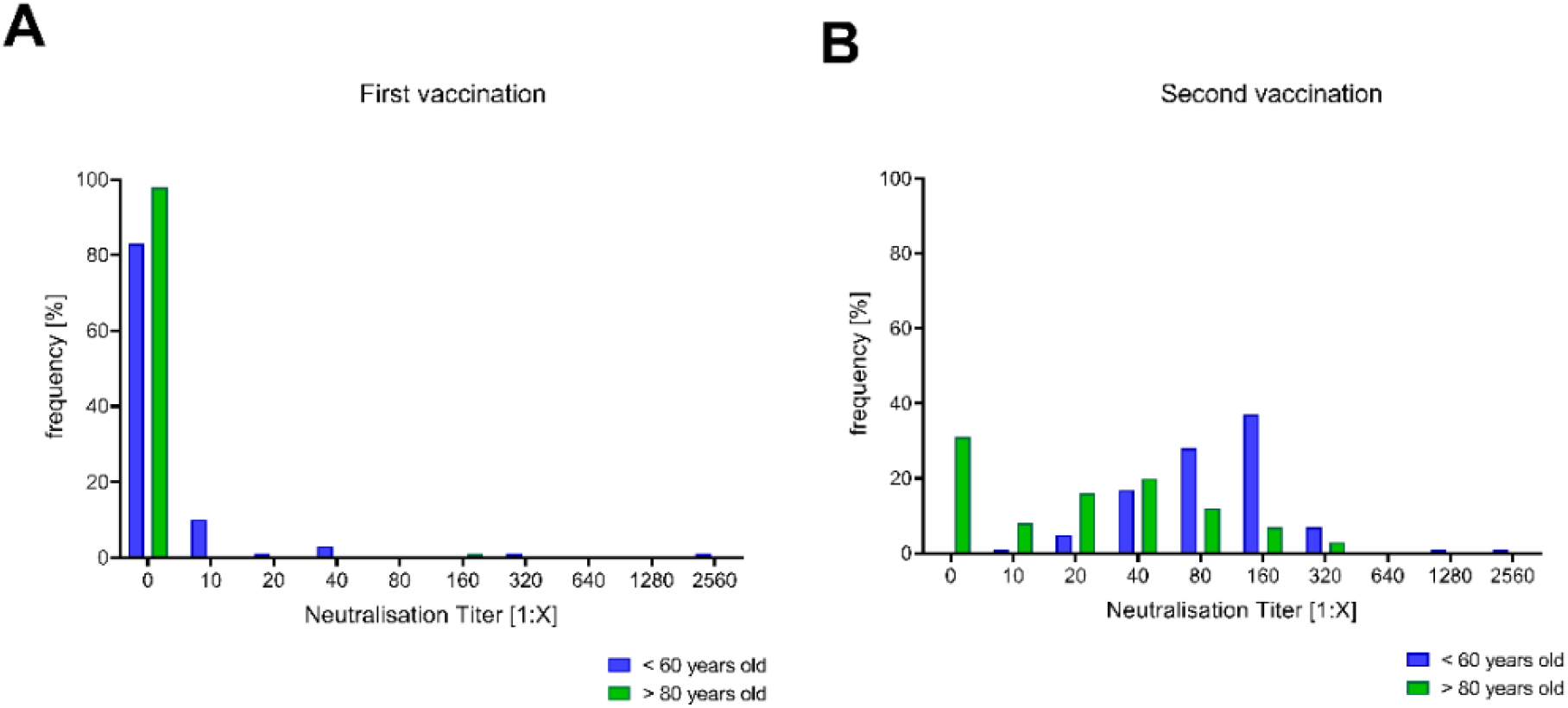
Neutralization antibody titers were determined as described in the methods section. The frequencies of individuals with a certain neutralizing antibody titer after the first vaccination (A) and the second vaccination (B) are shown.

After the second dose, a neutralization titer was attained by 97.8% of the younger vaccinees. In the elderly group, 68.7% showed titers ranging from 1:10 to 1:320. Remarkably, in 31.3% of the elderly vaccinees neutralizing antibodies were not detectable after the second vaccination, and thus, were potentially without seroprotection (Figure 2B).

### The severity of post-vaccination symptoms does not correlate with antibody response

To assess differences in post-vaccination symptoms between the age groups and to evaluate a potential correlation with antibody titers, medical questionnaires were completed at the two collection time points.

After the first vaccination, half of the younger cohort (51.6%) reported no symptoms, the remaining vaccinees recorded post-vaccination symptoms with a score ranging between 1 and 4. In turn, 93.9 % of elderly vaccinees reported no symptoms; the remaining 6.1% reported either one or two of the scored symptoms (Figure 3A).

**Figure 3.**
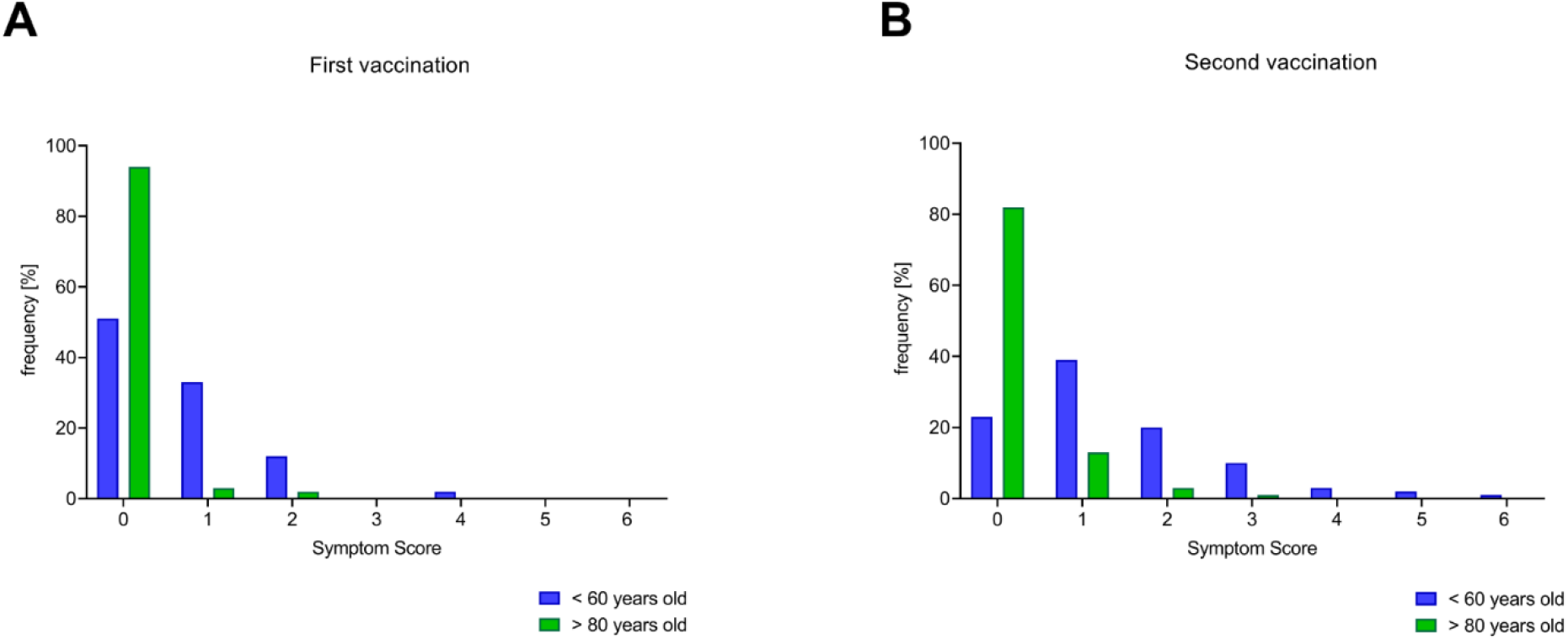
Symptom scores after first (A) and second (B) vaccination were determined as the sum of cumulative side effects using to the predefined categories (see method section).

After the second dose, only 25.8% of the younger vaccinees had no symptoms. While 38.7% of this group reported only one of the scored symptoms, 35.5% reported a combination of symptoms scoring between 2 and 6. Among the elderly, 83.1% reported no symptoms, and the remaining 16.9% of this group reported symptoms up to a score of 3 (Figure 3B). However, there was no general correlation between vaccination-induced SARS-CoV-2 spike specific IgG production and the presence or absence of individual symptom reports.

## Discussion

The SARS-CoV-2 pandemic has led to the development of various vaccines and vaccine strategies, which have been made available to the public by either emergency use designation or conditional marketing authorization. Inevitably, data on populations that are difficult to enroll including immunocompromised or cohorts <16 years or >80 years who might show reduced vaccine reactiveness are limited. The main goal of this real-life study was to investigate the efficacy of the current vaccination strategy in the most vulnerable group of vaccinees (>80 years old) compared to those younger than 60 years. We compared the induction of immune responses in these two age groups after the first and second BNT162b2 COVID-19 vaccination by measuring vaccine-induced SARS-CoV-2 spike specific IgG and SARS-CoV-2 neutralizing antibodies. While the majority of both young and elderly vaccinees raised IgG responses after their second vaccination, the induction of ELISA-IgG and in particular neutralizing antibody levels were significantly lower in the elderly vaccinees.

The main differences between the two groups of vaccinated individuals are likely a consequence of immunosenescence, which describes the phenomenon of reduced adaptive immune responses e.g. antibody responses in the elderly [23]. It is well described that elderly individuals not only have higher rates of morbidity due to infection but also respond less to vaccination [24–26], mainly due to a decline in cellular as well as humoral immunity.

The notion that humoral vaccination responses are impaired with increasing age is well depicted in our cohort, as the mean titer of SARS-CoV-2 spike specific IgG remains 2.8-fold lower after the second vaccination for the elderly group of vaccinees compared to the younger cohort (Figure 1B). Additionally, a general intra-and inter-group trend in negative correlation between age and IgG titer is visible after both vaccinations (Figure 1C/1D). More importantly, a similar age-dependent trend can be seen for SARS-CoV-2 specific neutralizing antibody titers: While neutralization antibody titers were attained by 97.8% of the younger vaccinees, 31.3% of the elderly remained without neutralization antibody titers after the second vaccination (Figure 2B).

The lack of neutralizing antibody responses in about one-third of the elderly group raises the questions whether the effectiveness of vaccine-induced immune protection may be transferred to this population without explicit testing. Especially since neutralizing, antibody levels correlate with protection against many viruses including SARS-CoV-2 in humans [27, 28] and recent data suggest that high neutralizing titers are particularly important for protection against novel circulating SARS-CoV-2 variants confering immune escape [29–31].

Although it is well known that the response to primary vaccination is weaker in the elderly [24, 32], it is remarkable that this observation also expands to the younger cohort. While there are reports that high neutralization titers were attained after receiving both doses of the Biontech/Pfizer Comirnaty (BNT162b2) vaccinations [18, 33, 34], which is in line with our data for the younger cohort, there is a lack of information on the antibody responses after the first vaccination. The latter gains particular importance since different vaccination schedules for the same vaccines have been adopted in several countries. These include a delay of the second vaccination, as implemented by the UK or Israel, to allow for the initial primary vaccination to a larger proportion of the population, a strategy which is controversially discussed [35, 36]. The observation that single-dose vaccinees broadly lacked neutralizing antibody responses quickly raises the question, whether these individuals might still acquire infections and may transmit the disease while remaining asymptomatic. This assumption is supported by recent results of a large Israeli study which reports a 46% effectiveness in preventing a documented infection 14 to 20 days after the first dose, the BNT162b2 vaccine [37].

While this study was in progress, the first promising reports on the experience with COVID vaccination came from Israel and Scotland [38]. It appears that even after the first SARS-CoV-2 vaccination, a significant decrease in hospitalizations is seen in the overall vaccinated population but also the >80 year old group. However, it is not yet clear how long this protective effect of vaccination lasts. Our data presented here suggests that it might be necessary to define strategies to overcome age-related limitations for COVID-19 vaccination. Moderna has recently demonstrated an increased immune response determined by higher binding and neutralizing antibody titers by increasing the dose of the second vaccination from 25 µl to 100 µl [39]. Strategies to enhance immunogenicity such as the use of adjuvants, application of increased amounts or multiple doses of the same vaccine, or the combination of different vaccines for a heterologous prime/boost should be rapidly tested and implemented in COVID-19 vaccination protocols. Furthermore, since the majority of vaccinees did not obtain neutralizing antibody titers after the first vaccination, we suggest that postponing a second vaccination with this vaccine is neither advisable for younger nor elderly populations.

This study provides insight into age-dependent limitations of immune responses elicited after the first and second dose of the BNT162b2 vaccine. By comparing similar-sized cohorts of vaccinees aged < 60 years and > 80 years, we found that more than 30% of elderly vaccinees did not attain neutralizing antibody responses after their second vaccination. Nevertheless, recent studies show that even after the first vaccination, severe courses of COVID-19 are attenuated. The elderly population is prioritized by many vaccination schedules, despite the fact that this age group is underrepresented in previous studies, and hence, there is still a lack of data concerning the induced immune response after both, first and second vaccination in this population.

## Data Availability

There is no supplemental data available.

## Notes

## Acknowledgments

The authors thank Dr. Anna Seelentag (Sozial Betriebe Köln, SBK) for excellent organization and support, Gabriele Patzke (SBK) for the opportunity to conduct this study, Dr. Irene Spiertz-Schmidt, Mrs. Mavinga and the caregivers at SBK for their support at the blood collection days. We would like to thank all volunteers at the SBK nursing home. We thank all members of the Virology department.

## Author contributions

Conceptualization: HS, OA, MA, LM, Formal analysis: OA, WM, Investigation: LM, MA, WM, ID, LW, RG, JP, JH, AR, DR, OA, HS, Writing – original draft preparation: HS, OA, MA, LM, Writing – review and editing: LM, MA, WM, ID, LW, RG, JP, JH, AR, DR, PNO, RR, SH, AW, CM, RG, JT, OA, HS, Supervision: HS, OA, MA, LM

